# Neural processing of social reciprocity in autism

**DOI:** 10.1101/2024.01.31.24302051

**Authors:** A. M. Bierlich, I. S. Plank, N. T. Scheel, D. Keeser, C. M. Falter-Wagner

**Affiliations:** Department of Psychiatry and Psychotherapy, LMU University Hospital, LMU Munich Nussbaumstrasse 7, 80336, Munich, Germany; NeuroImaging Core Unit Munich (NICUM), LMU University Hospital, LMU Munich Nussbaumstrasse 7, 80336, Munich, Germany

**Keywords:** autism, social interactions, reciprocity, interpersonal synchrony, fMRI

## Abstract

Social reciprocity and interpersonal synchrony implicitly mediate social interactions to facilitate natural exchanges. These processes are altered in autism, but it is unclear how such alterations manifest at the neural level during social interaction processing. Using task-based fMRI, we investigated the neural correlates of interpersonal synchrony during basic reciprocal interactions in a preregistered study. Participants communicated with a virtual partner by sending visual signals. Analyses showed comparable activation patterns and experienced synchrony ratings between autistic and non-autistic participants, as well as between interactions with virtual partners who had high or low synchronous responses. An exploratory whole brain analysis for the effect of task revealed significant activation of the right inferior frontal gyrus (IFG) and left anterior inferior parietal lobe (IPL); areas associated with cognitive control, temporal coordination, and action observation. This activation was independent of the virtual partner’s response synchrony and was similar for autistic and non-autistic participants. These results provide an initial look into the neural basis of processing social reciprocity in autism, particularly when individuals are part of an interaction, and hint that the neural processing of social reciprocity may be spared in autism when their partners’ behavior is predictable.

## 1. Introduction

Social interactions are complex situations, in which nuanced features govern behaviors across multiple modalities. Alterations in social reciprocity are a defining feature of Autism Spectrum Disorder (World Health Organization, 2016), whereby autistic individuals often struggle to achieve smooth social interactions. The multi-faceted nature of these atypical social interactions has been broadly investigated (Fusaroli et al., 2017; McNaughton & Redcay, 2020; Philip et al., 2012; Velikonja et al., 2019), with notable focus on alterations in higher-order cognitive processes, such as emotion processing and Theory of Mind, as key reasons underlying social interaction difficulties in autism. However, less research has focused on fundamental features that constitute and regulate the interactional experience when one is part of an interaction.

The quality of social interactions is highly influenced by interpersonal synchrony, which is the temporal coordination of behaviors between interactants (Bowsher-Murray et al., 2022; Hoehl et al., 2021). Interpersonal synchrony has been shown to boost rapport and facilitate social affiliation between interactants (Hove & Risen, 2009; Lakens & Stel, 2011; Miles et al., 2009). Growing evidence shows that interpersonal synchrony is attenuated in relation to autism (Georgescu et al., 2020; Koehler et al., 2021; McNaughton & Redcay, 2020; Zampella et al., 2020). For example, dyads including an autistic individual (or dyads of two individuals with autism) tend to show less synchronous behavior than dyads including two non-autistic individuals (Georgescu et al., 2020). Such insights into the differentiation of interpersonal synchrony have sparked investigation into its relevance for the diagnostic classification of autism (Georgescu et al., 2019; Koehler et al., 2021, 2022; Koehler & Falter-Wagner, 2023; Plank et al., 2023). However, the mechanistic root underlying altered interpersonal synchrony, as a function of atypical reciprocity, in autism has not yet been explored.

Joint action research broadly addresses how such behaviors are predicted, perceived, integrated, and coordinated to jointly achieve a goal, wherein autistic individuals may struggle with or use different mechanisms (Cerullo et al., 2021). Temporal cues seem to be especially useful for coordination during joint interactions. Using a drum tapping paradigm, Yoo and Kim (2018) found that rhythmic cueing was beneficial for autistic individuals to synchronize their own drumming with a partner. If temporal information, *not elicited* by the partner, can enhance coordination for autistic individuals, then temporal information, *indeed elicited* by a partner, should also enhance coordination but may be differentially processed by autistic individuals. As such, differences in the perception of timing of other’s behaviors may account for attenuated interpersonal synchrony and, in turn, atypical social reciprocity.

Some alterations in time perception have been shown in autism (Allman, 2011; Allman & Falter, 2015; Casassus et al., 2019). Although, these differences are dependent on the specific task at hand; for instance, studies using simultaneity judgment paradigms have found enhanced temporal resolution in autism on the behavioral (Falter et al., 2012) and neural level (Falter et al., 2013; Menassa et al., 2018). Meanwhile, other studies have reported no differences between autistic and non-autistic individuals in different temporal processing domains, such as duration discrimination, clock timing, and relative timing (e.g., Isaksson et al., 2018; Poole et al., 2022; for a comprehensive review, see Casassus et al., 2019). Although interpersonal synchrony and reciprocal timing arguably require accurate perception of an interactant’s behavior, it remains unclear whether and to what extent temporal processing is altered in autism and how it impacts social reciprocity.

Addressing such fundamental features of social exchange presents a challenge for the experimental design to maintain the social nature of the interaction while still isolating the modulatory aspects of interpersonal synchrony and reciprocity. Few studies have approached this caveat (e.g., Bowsher-Murray et al., 2023; Cacioppo et al., 2014; Cirelli et al., 2014). In a sample of adults without a diagnosis, Cacioppo et al. (2014) investigated the neural processing of interpersonal synchrony using a basic reciprocal interaction with a virtual partner, which was modulated by participants’ own responses and computer-driven response latencies. Participants communicated with the virtual partner by sending signals via button presses. The authors found higher ratings of experienced synchrony, as well as increased neural activation in the ventromedial prefrontal cortex (vmPFC), parahippocampal gyrus, and inferior parietal lobe (IPL), for virtual partners whose responses showed greater consistency and, thus, were more synchronous with their own responses. This paradigm lends itself to an investigation of reciprocal interaction in autism, stripping away highly complex social information to capture the fundamental processes related to synchrony in reciprocal interactions.

Therefore, the current preregistered study dissected the underlying processes of basic reciprocal interactions and investigated whether they might contribute to social interaction difficulties apparent in autism, using the same paradigm as Cacioppo et al. (2014). We investigated neural activation patterns and subjective ratings of interpersonal synchrony from autistic and non-autistic participants when they engaged in a nonverbal, reciprocal interaction with a virtual partner. Behaviorally, we expected that participants’ ratings of experienced synchrony and rapport with the virtual partner would increase as a function of synchrony. Likewise, ratings were expected to be lower in autism. At the neural level, we expected that neural activation would differ between autistic and non-autistic individuals as well as between more and less synchronous conditions, as defined by the immediacy and regularity of the virtual partner’s responses. Specifically, we expect to observe differences in neural activation in areas associated with cognitive control, temporal coordination, as well as inferring others’ actions and intentions (Cacioppo et al., 2014; Sakaiya et al., 2013).

## 2. Methods

### 2.1. Participants

We recruited 33 autistic and 29 non-autistic individuals from the LMU Munich autism outpatient unit, regional network partners, and through local channels (e.g., university email lists). The sample size of at least 25 analyzed participants per group was based on a power analysis conducted in G*Power 3.1 (Faul et al., 2009), considering an effect size of 0.31 (partial eta^2^ = 0.09, (Cacioppo et al., 2014)), power of 0.99, and an alpha level 0.05 as based on a repeated measure, within-between interaction ANOVA.

Autistic participants had a confirmed ICD-10 diagnosis of F84.5, F84.0, or F84.1 (World Health Organization, 2016). All diagnoses of autism were given based on diagnostic procedures adhering to the German health guideline for diagnostics of autism (AWMF, 2016), as confirmed by medical records. Non-autistic participants had no psychiatric diagnoses as per self-report. Exclusion criteria were age under 18 years or older than 60 years, neurological diagnoses, metal or cochlear implants, pacemakers, or an IQ below 70. All participants had normal or corrected-to-normal vision. Verbal and nonverbal IQ were assessed using the Mehrfachwahl-Wortschatz-Intelligenztest (MWT-B; Multiple choice vocabulary test; Lehrl et al., 1999) and the Culture Fair Test (CFT-20-R; Weiß, 2006), respectively.

The first three tested non-autistic individuals were omitted after minor adjustments to the task were made, one autistic individual was not included due to an IQ below 70, and data of six individuals (one non-autistic, five autistic) was excluded from analysis due to excessive head movement (> 3mm) while in the scanner. The final sample analyzed consisted of 27 autistic (9 identified as female, 18 as male) and 25 non-autistic individuals (8 identified as female, 17 as male). Groups were matched for age and IQ but differed in autism-like traits and motor coordination (Table 1), as measured by the German translations of the Autism Spectrum Quotient (AQ; Baron-Cohen et al., 2001) and Adult Dyspraxia Checklist (ADC; Kirby et al., 2010).

**Table 1.**
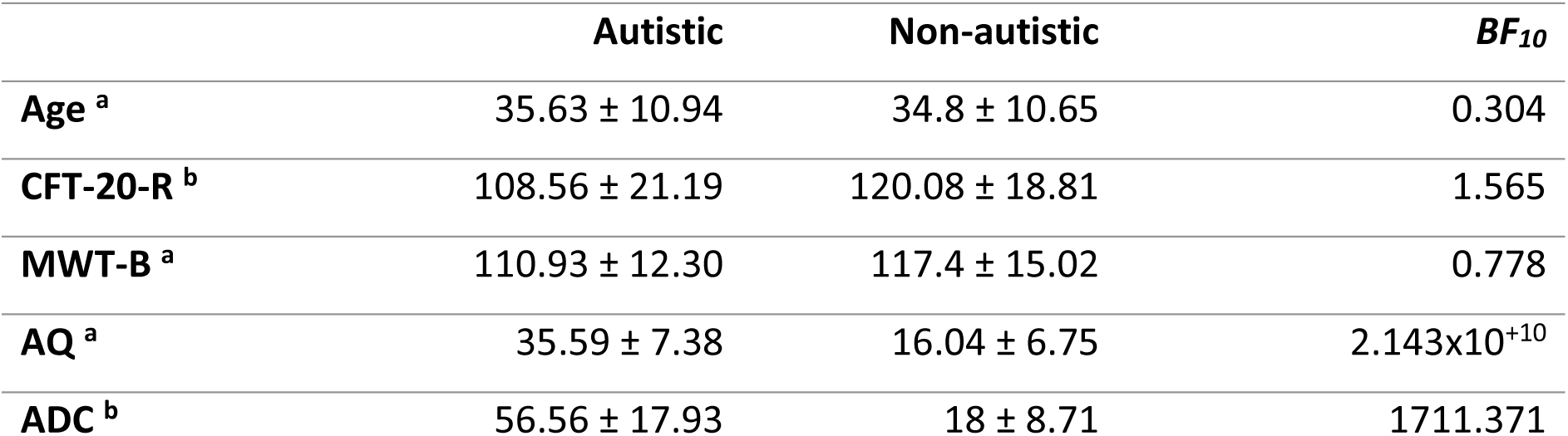
Means and standard deviations are reported for each group, as well as the Bayes Factor from a Bayesian Mann-Whitney U test (^a^) or Bayesian independent samples t-test (^b^).

|  | **Autistic** | **Non-autistic** | ***BF*<sub>10</sub>** |
| --- | --- | --- | --- |
| **Age** <sup>a</sup> | 35.63 ± 10.94 | 34.8 ± 10.65 | 0.304 |
| **CFT-20-R** <sup>b</sup> | 108.56 ± 21.19 | 120.08 ± 18.81 | 1.565 |
| **MWT-B** <sup>a</sup> | 110.93 ± 12.30 | 117.4 ± 15.02 | 0.778 |
| **AQ** <sup>a</sup> | 35.59 ± 7.38 | 16.04 ± 6.75 | 2.143x10<sup>+10</sup> |
| **ADC** <sup>b</sup> | 56.56 ± 17.93 | 18 ± 8.71 | 1711.371 |

Participants provided informed consent, in accordance with the Declaration of Helsinki (World Medical Association, 2013), to participate in the study. This study was approved by the ethics committee of the Medical Faculty at LMU Munich (No. 20-1050). A preregistration can be found with the OSF project: https://osf.io/cw7n4.

### 2.2. Experimental Design

A nonverbal, basic, reciprocal interaction paradigm was employed (Cacioppo et al., 2014). Participants communicated with a virtual partner by sending signals via button presses. The participant and virtual partner were visually represented by shape avatars, wherein the shape would change when a signal was sent (Fig 1). During their turn, participants were instructed to send a signal roughly once per second, to which the virtual partner would respond. In actuality, the virtual partner’s responses were computer-generated and manipulated by the signals’ mean *latency* and *dispersion* (range from a uniform distribution). This comprised two features of reciprocity that are important for emulating synchronous behaviors (Bowsher-Murray et al., 2023). Each feature consisted of a low and high condition, resulting in four virtual partners: low latency-low dispersion (120 ± 10 ms), low latency-high dispersion (120 ± 110 ms), high latency-low dispersion (220 ± 10 ms), high latency-high dispersion (220 ± 110 ms).

**Fig 1.**
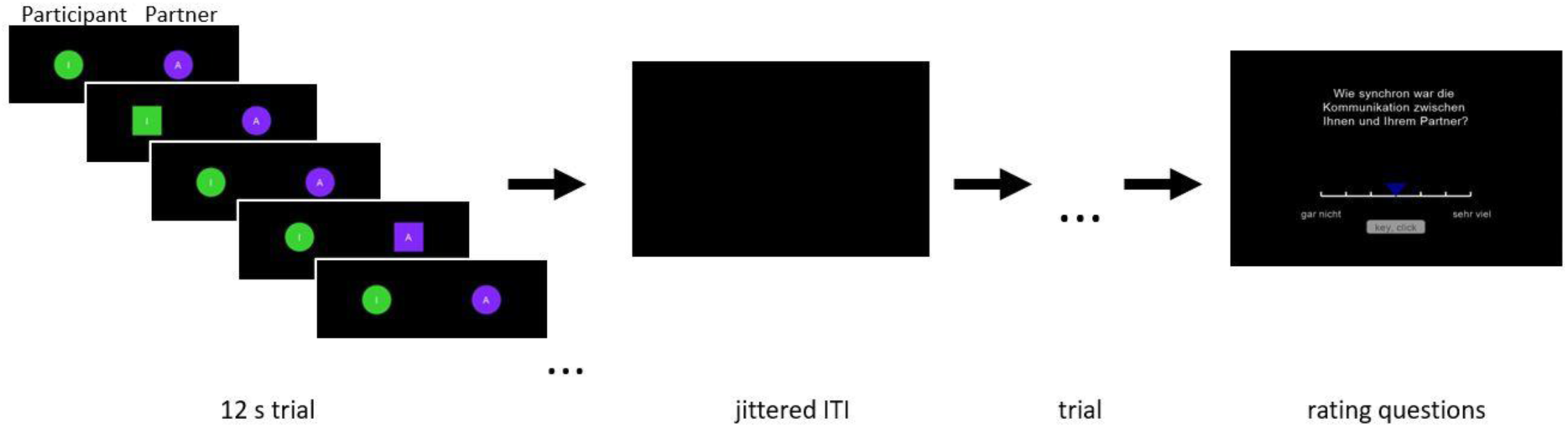
Depiction of the sequence for each block beginning with the communication sequence within each 12-second trial and ending with an example of a rating question in German.

Each virtual partner was defined by one block of eight 12-second trials that were interleaved by an intertrial interval to allow the neural activation to return to baseline. Each trial had a different response latency within the defined response range (e.g., 120 ± 10 ms). The virtual partner’s response latency was pseudo-randomly selected within each response range for each participant. Intertrial intervals (ITIs) were determined using a uniform distribution in the neurodesign library (Durnez et al., 2017), resulting in ITIs of 4.4, 4.9, 5.2, and 5.5 seconds. Of these ITIs, one was randomly chosen for each trial. After each block, participants were asked to rate the interaction with their virtual partner in seven questions. Six questions asked about the rapport felt with the virtual partner and one question asked about experienced synchrony (Supplementary Information S1). The questions were always asked in the same order, and the question regarding experienced synchrony was always placed fourth. These questions were translated from English (Cacioppo et al., 2014) into German, as the data was collected from a German-speaking sample. Participants responded on a 7-point Likert scale anchoring 1 = ‘not at all’ to 7 = ‘very much’. The slider starting position was anchored at the fourth marker (i.e. in the middle). Participants had eight seconds to confirm their rating, or ‘None’ would be recorded. Block order was pseudo-randomly presented across participants. An example sequence is depicted in Fig 1. Stimuli were presented using Psychopy (Peirce et al., 2019), with Python 3.8 and Anaconda Navigator (Version 2.0.4).

### 2.3. Procedure

In a training round outside of the scanner, participants familiarized themselves with the task, response boxes, and rating scales. One experimenter instructed the task, while the other experimenter acted as the interaction partner at a mock station during the training. Two confederates were also present. The participant was told that these four individuals will be their interaction partners during the task. This cover story was intended to boost interaction believability. Subsequently, participants performed the task in the scanner. On the same day, participants performed additional tasks and questionnaires of an unrelated study that will be presented elsewhere (see the preregistration for details).

Following the testing session, participants were asked two questions about the quality of the interaction (Supplementary Information S2), as well as debriefed about the intent behind the study. The rating questions were based on a 10-point scales. The quality of the interaction was rated from 1 = ‘very bad’ to 10 = ‘very good’, and how much it felt like an interaction was rated from 1 = ‘very little’ to 10 = ‘very much’. An experimenter observationally classified participants’ reactions to the interaction task: (i) did not at all suspect that they were interacting with a computer (n = 22), (ii) stated suspicion of interacting with a computer only after it was specifically mentioned by the experimenter (n = 18), and (iii) immediately offered their suspicions of interacting with a computer (n = 9). Classifications are missing for three participants because the debriefing procedure was further optimized.

### 2.4. Behavioral Preprocessing and Analysis

Preprocessing was conducted in RStudio 2023.6.0 (Posit Team, 2023), using R 4.2.2 (R Core Team, 2022). Behavioral analyses were conducted in JASP 0.17.2.1 (JASP Team, 2023).

Fifteen rating responses (5 experienced synchrony, 10 rapport-related) were recorded as “None” (Supplementary Information S3). Using Cronbach’s alpha, the six rapport questions demonstrated high internal consistency across condition and group (α = .937), so they were aggregated into a mean composite rapport rating. To test the behavioral hypotheses, Bayesian mixed ANOVAs were conducted, including diagnostic group (autistic/non-autistic), latency (low/high), and dispersion (low/high) as factors.

In an exploratory analysis, the individual tapping behaviors (i.e., sending a signal to the virtual partner) were also computed to assess the total frequency for each condition, as well as participants’ mean response latency. For two participants, tapping behaviors were missing for the first trial due to participant error. Like the response ratings, tapping behaviors were also analyzed using a Bayesian mixed ANOVA including diagnostic group (autistic/non-autistic), latency (low/high), and dispersion (low/high) as factors. Bayes factors were interpreted according to the Jeffrey’s scheme (Goss-Sampson, 2020).

### 2.5. fMRI Data Acquisition

Neuroimaging data were collected at the Neuroimaging Core Unit Munich (NICUM) at the LMU University Hospital Munich using a 3 Tesla Siemens MRI scanner (Siemens Magnetom Prisma, Siemens Medical Solutions, Erlangen, Germany). First, structural T1-weighted scans were collected (176 slices; voxel size = 1 mm^3^; TR = 2250 ms; FOV = 256 mm) followed by field maps (32 slices; voxel size = 3 mm^3^; TR = 8000 ms; TE = 66 ms; FOV = 210 mm). Then, T2-weighted echo-planar imaging (EPI) sequences measured brain activation during the fMRI tasks (564 volumes; 32 slices; voxel size = 3 mm^3^; TR = 2066 s; TE = 30 ms; FOV = 210 mm).

### 2.6. fMRI Preprocessing

NIFTI conversion was conducted using dcm2bids 2.1.6 (Bedetti et al., 2021). fMRI pre-processing was performed using fMRIPrep 22.1.1 (Esteban et al., 2019). The automatically generated template from fMRIPrep details the pre-processing pipeline and is reported in Supplementary Information S4. The T1-weighted anatomical scans were used as references after undergoing correction for non-uniformity, skull-stripping, brain tissue segmentation, surface reconstruction (FreeSurfer 7.2.0; Dale et al., 1999), spatial normalization, and registration to MNI152 standard space templates. Each EPI scan underwent field map correction, slice time correction, and co-registration to the T1-weighted reference. Spatial smoothing (Gaussian kernel 6mm FWHM) and motion artifact removal using ICA-AROMA (Pruim et al., 2015) were performed. Participants were omitted if their head motion exceeded more than 1 voxel (3 mm) for any of the three translational head motion parameters. The resampled brain mask was applied to the preprocessed EPI scans using fslmaths (Smith et al., 2004).

### 2.7. fMRI Analysis

fMRI post-data analysis was performed using FSL FEAT (Smith et al., 2004). At the subject-level, a general linear model, including the four partner interaction conditions and the communication frequency as a parametric modulator, was constructed. Four differential contrasts were created to assess the activation with respect to the manipulated latency (low > high, high > low) and dispersion (low > high, high > low). Two additional differential contrasts were included to assess the interaction effect of latency and dispersion. A final contrast of the pooled conditions was included to capture the activation across the task. At the group-level, one-sample t-tests compared the differential contrasts of latency and dispersion in a pooled sample, and unpaired t-tests compared the autistic and non-autistic groups for each of the differential contrasts. A region of interest (ROI) analysis was employed to test each hypothesis (Supplementary Information S5). ROIs were anatomically defined using Marina in a single mask (Walter, 2003), including the medial superior frontal gyrus (left/right), anterior cingulate cortex (left/right), amygdala (left), parahippocampal gyrus (left), inferior parietal lobe (left), anterior gyrus (left), and the supramarginal gyrus (left). These regions captured the peak voxel activation reported by Cacioppo et al. (2014). Additionally, an exploratory whole brain approach assessed activation outside of the defined ROIs for each contrast. Exploratory sub-analyses were conducted, including (i) a replication analysis of the non-autistic participants using a parametric approach at the whole brain level, and (ii) an analysis considering the debrief assessment of interaction believability. Hypothesized and replication analyses were validated using SPM12 (Wellcome Department of Imaging Neuroscience, University College London, UK, 2014). They are reported in Supplementary Information S6. Group-level analyses were conducted using a non-parametric approach with FSL randomise (TFCE (Smith & Nichols, 2009), 5000 permutations, *p* < .05) to better account for false positives (Eklund et al., 2016). Identified brain regions are specified from the Automated Anatomical Labeling from MRIcroGL (Rorden & Brett, 2000). The quality metrics (Esteban et al., 2017) as well as exploratory analyses of the structural T1w data (processed using NAMNIs; Karali et al., 2021) between autistic and non-autistic participants are reported in Supplementary Information S7 and S8.

## 3. Results

### 3.1. Behavioral Results

#### 3.1.1. Subjective Ratings

The experienced synchrony ratings (Fig 2a) were best explained by the null model (Supplementary Information S9). The second-best model included group but was only half as likely as the null model (*BF_10_* = 0.509); thus, rejecting our behavioral hypotheses regarding differences between groups and conditions with respect to experienced synchrony ratings.

**Fig 2.**
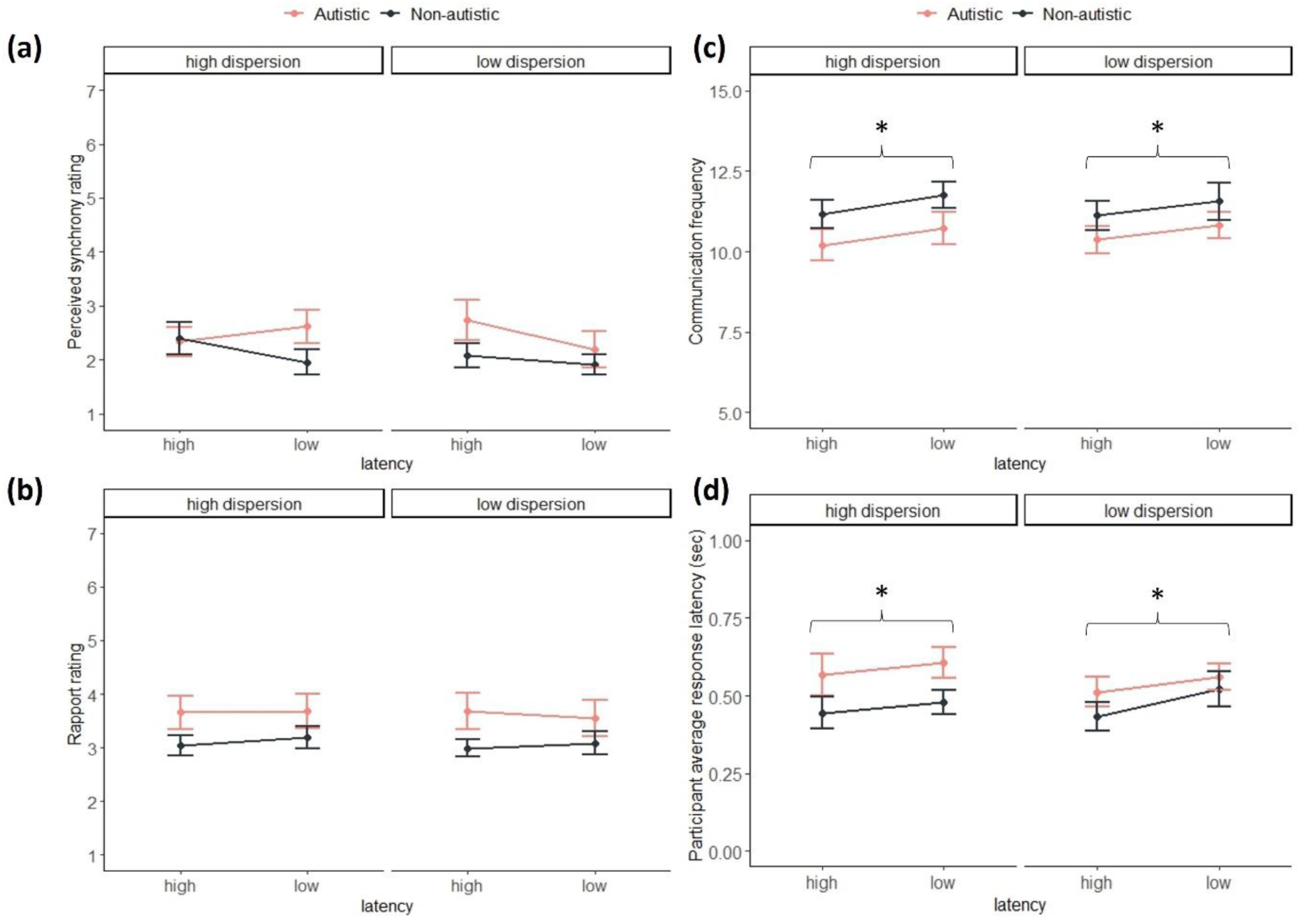
(A) Mean and standard error of the experienced synchrony ratings for the low and high latency and dispersion conditions for each group. Based on the scale: 1 (‘not at all’) to 7 (‘very much’). (B) Mean and standard error of rapport ratings for the low and high latency and dispersion conditions for each group. Based on the scale: 1 (‘not at all’) to 7 (‘very much’). (C) Mean and standard error of the communication frequency for the low and high latency conditions for each group. (D) Mean and standard error of participants’ average response latencies for the low and high latency conditions for each group. Stars indicate a significant effect of latency.

When assessing the rapport ratings (Fig 2b), the results identified that a model including group best fits the data (*BF_10_* = 1.040), although it is only marginally more likely than the null model. This hints at anecdotal evidence in favor of group differences, as driven by autistic participants reporting greater rapport with the virtual partners than non-autistic participants. However, the evidence is not strong enough to support our behavioral hypotheses regarding differences between diagnostic groups and conditions with respect to rapport ratings.

#### 3.1.2. Tapping Behaviors

As a sanity check, we compared participants’ tapping behaviors to confirm that they performed as instructed. When assessing participants’ communication frequency (i.e., how often they sent a signal to the virtual partner), the results yielded very strong evidence that the model that best explained the data only included the factor latency of the virtual partner’s response (*BF_10_* = 40.560). This was further supported by the inclusion Bayes Factor with very strong evidence in favor of including the factor latency in the model (*BF_incl_* = 39.734) and indicated that participants sent more signals to the virtual partner when the partner’s response latency was faster (Fig 2c). Similarly, when assessing participants’ average response latencies, the results yielded strong evidence that the latency condition of the virtual partner best explained the data (*BF_10_* = 14.886). This was supported by the inclusion Bayes Factor with strong evidence in favor of including the factor latency (*BF_incl_* = 15.288). Participants’ average response latencies were faster when the partner’s responses were faster (Fig 2d).

### 3.2. fMRI Results

In the ROI analysis, neural activation of the pooled sample was comparable between the respective contrasts for dispersion (low > high; high > low) and latency (low > high; high > low), as well as their interaction, rejecting our first fMRI hypothesis regarding differences between synchrony conditions. Likewise, the autistic and non-autistic groups demonstrated similar activation in the ROIs for each of the contrasts; thus, rejecting our second and third fMRI hypotheses regarding differences between diagnostic groups and the interaction effect between diagnostic group and synchrony conditions.

To further explore possible activation outside of our predefined ROIs, a whole brain approach was used to assess the contrasts of latency (low > high; high > low) and dispersion (low > high; high > low) in a pooled sample and between diagnostic groups. Neural activation was comparable between conditions in pooled sample analyses, as well as between diagnostic groups in each of the contrasts.

Moreover, we assessed the effect of the task when the conditions and diagnostic groups were pooled using a one-sample t-test, wherein five clusters remained after correction (Table 2; Fig 3a). Notably, participants elicited significant activation in the right inferior frontal gyrus (IFG), in addition to the expected motor and visual cortices. Activation in the left motor cortex extended into the anterior region of the IPL (Fig 3b). Interestingly, when exploring the task effect between diagnostic groups, the autistic and non-autistic individuals elicited comparable task activation.

**Fig 3.**
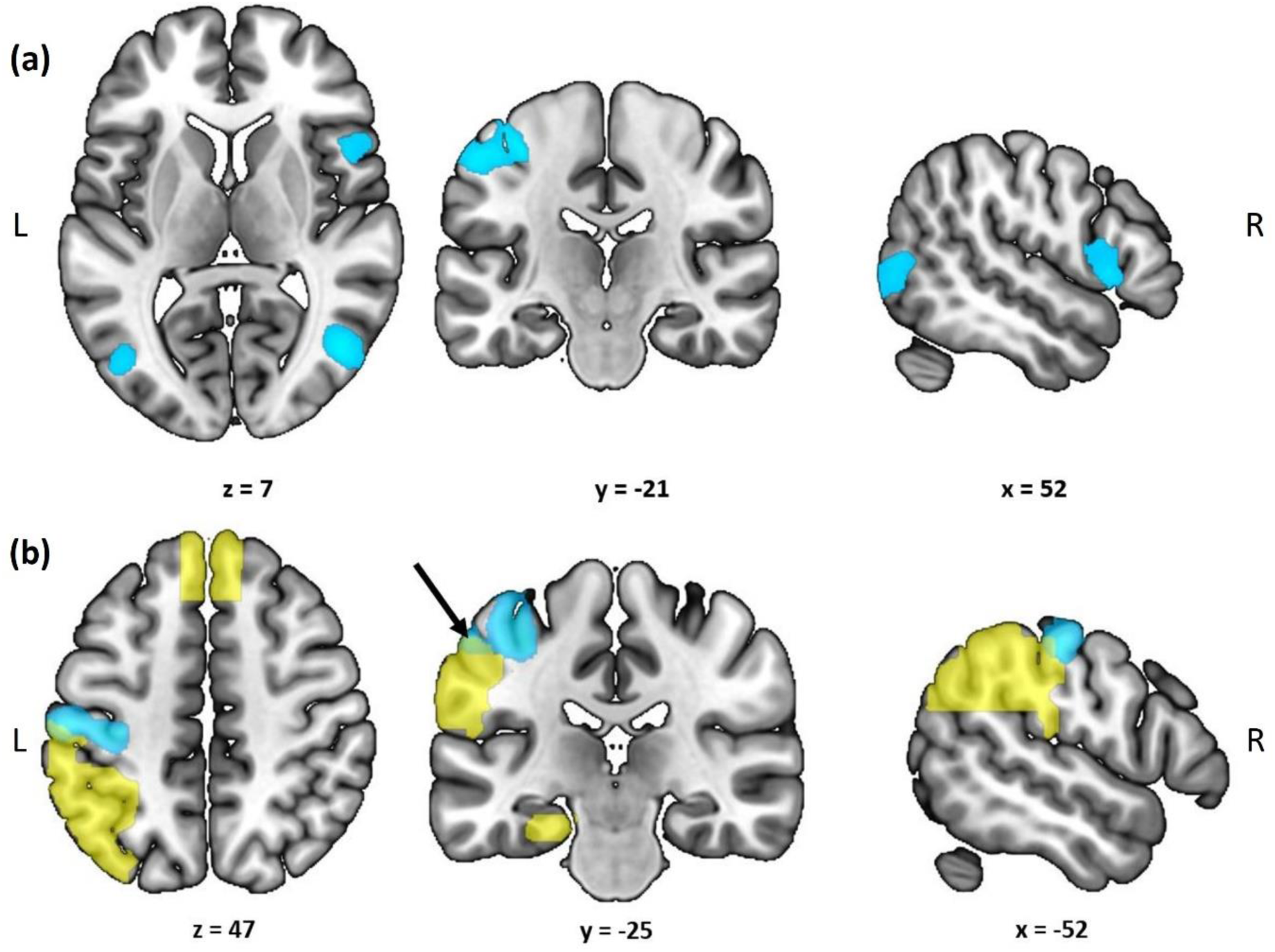
(A) Results of the whole brain analysis of the entire sample pooled across conditions. A t-contrast is shown highlighting surviving clusters (blue) from a non-parametric, TFCE approach considering p < .05. Participants showed significant activation in the right inferior frontal gyrus, bilateral inferior lateral occipital cortex, left motor cortex (precentral and postcentral gyri) extending into the anterior inferior parietal lobe, and the supplementary motor area (not pictured). (B) Results of the whole brain analysis of the entire sample pooled across conditions. A t-contrast (same as in Fig 2a) is shown highlighting surviving clusters (blue) from a non-parametric, TFCE approach considering p < .05. The hypothesized ROI mask is overlaid (yellow), highlighting parts of the surviving cluster that fall within the IPL ROI (green). Surviving clusters for the effect of task overlaid over an uncorrected t statistic map in Supplementary Information S10.

**Table 2.** Results of the whole brain analysis of the entire sample pooled across conditions. All significant clusters are reported from a non-parametric, TFCE approach considering p < .05. There were no significant clusters for the differential contrasts (latency, dispersion) or between diagnostic groups (autistic, non-autistic). Coordinates are reported in MNI space. BA: Brodmann Area; H: Hemisphere; L: Left; R: Right

| Region | BA | H | Cluster Size | t-value | x | y | z |
| --- | --- | --- | --- | --- | --- | --- | --- |
| **Postcentral Gyrus** | 4 | L | 1312 | 9.61 | -34 | -24 | 50 |
| Inferior Parietal Lobe | 1 | L |  | 8.18 | -54 | -22 | 48 |
| Precentral Gyrus | 4 | L |  | 7.73 | -40 | -18 | 58 |
| Supramarginal Gyrus | 40 | L |  | 6.87 | -55 | -21 | 46 |
| **Middle Temporal Gyrus** | 19 | R | 1303 | 9.13 | 44 | -70 | 4 |
| Middle Occipital Gyrus | 19 | R |  | 8.89 | 42 | -70 | 4 |
| Inferior Occipital Gyrus | 19 | R |  | 8.69 | 44 | -76 | -2 |
| Inferior Temporal Gyrus | 19 | R |  | 7.73 | 43 | -70 | -2 |
| **Supplementary Motor Area** | 6 | R | 157 | 8.09 | 6 | 0 | 70 |
| Supplementary Motor Area | 6 | L |  | 5.12 | -6 | -4 | 68 |
| **Inferior Frontal Gyrus, par. op.** | 44 | R | 383 | 7.56 | 54 | 16 | 2 |
| Inferior Frontal Gyrus, tri. | 45 | R |  | 6.30 | 54 | 20 | 2 |
| **Middle Occipital Gyrus** | 19 | L | 232 | 7.35 | -42 | -72 | 0 |
| Inferior Occipital Gyrus | 19 | L |  | 6.12 | -42 | -74 | -2 |

## 4. Discussion

Despite extensive research on social cognition in autism (e.g., Oberman & Ramachandran, 2007; Sato & Uono, 2019; Velikonja et al., 2019), little is known about the neural basis of how social interactions are processed, especially regarding implicit features like reciprocity and interpersonal synchrony. In the present preregistered study, we used task-based fMRI to investigate the neural correlates of interpersonal synchrony processing using a basic reciprocal interaction task, whereby participants communicated with a virtual partner. The virtual partner’s response was computer generated to emulate conditions that were more or less synchronous (i.e., constellations of low vs. high latency and dispersion) and based on participants’ own responses. When interacting with the virtual partners, autistic and non-autistic participants reported comparable experienced synchrony and rapport for low and high synchrony partners. At the neural level, we also found similar activation patterns for the synchronous and asynchronous virtual partners, as well as between autistic and non-autistic participants for each synchrony condition. Furthermore, whole brain analyses demonstrated distinct regions activated when engaging in reciprocal interactions across synchrony conditions. Importantly, autistic individuals engaged the same brain regions when they were involved in a reciprocal interaction as non-autistic individuals.

These results could indicate that autistic and non-autistic participants process and experience synchrony and rapport in a similar way, as evident by the similar neural and behavioral responses. Koehne et al. (2016) also found that autistic and non-autistic individuals reported similar experienced synchrony, when investigating its relationship to empathy in a behavioral leader-follower task. Likewise, Bowsher-Murray et al. (2023) recently investigated how interpersonal synchrony influences affiliation in children during a pseudo-interaction with a virtual partner, wherein they found comparable affiliation ratings for synchronous and asynchronous virtual partners. Another possible explanation for the results in the current study may stem from the simplistic social nature of the paradigm, requiring participants to engage with (i) a non-visible partner, in a (ii) repetitive activity, and requiring (iii) simple button presses. As such, the paradigm, although socially reciprocal, might not be demanding enough to elicit the difficulties with reciprocal social interactions that are symptomatic of autism. It is plausible that differences between autistic and non-autistic individuals may lie in the processing of more complex and nuanced social information. Further investigation is necessary to clarify the extent to which experiencing and perceiving interpersonal synchrony is intact in autism when (i) using a more socially engaging task (e.g., avatars rather than shapes, movement cues, or simulated video-based interactions), and (ii) adding increasingly complex social information (e.g., social cues, emotional states) to assess possible differences in processing more demanding reciprocal interactions and how they are influenced by interpersonal synchrony in autism.

The choice of a simplified paradigm in the current study was to investigate basic behavioral and neural processes of social interaction in autism as well as to allow for comparability with previous work in non-autistic adults that reported differentiated activation in the vmPFC as linked to experienced synchrony using the same paradigm (Cacioppo et al., 2014). However, in contrast to the previously reported findings, when investigating the neural correlates of interpersonal synchrony perception, we did not find differentiated activation for virtual partners with low and high synchronous responses. The lack of a differential neural effect is in line with participants reporting similar ratings of experienced synchrony and rapport. Importantly, the differential neural effect was also not replicable for non-autistic participants in the current study, despite sufficient power and a larger sample size compared to the original study as well as task replication. Thus, synchrony may not influence basic reciprocal interactions at the neural and behavioral levels; however, in light of previous research showing synchrony’s influence on behavior and perception (Hoehl et al., 2021; Mogan et al., 2017), it is possible that the manipulations of latency and dispersion were not extensive enough to convey synchronous and asynchronous behaviors. In line with this rationale, Koehne et al. (2016) used increased response latencies (500 and 750 ms) and larger dispersion (± 10% half of the nominal distribution of the mean latencies) between virtual partners, in which they found differences in subjective reports of experienced synchrony between more or less synchronous virtual partners. Thus, more apparent differences between synchrony manipulations may be necessary to elicit reliable differences in processing interpersonal synchrony.

We also assessed participants tapping behaviors to check whether they were influenced by synchrony manipulations of the virtual partners. Virtual partners’ mean response latencies, but not dispersion, influenced participants’ communication frequency and average response latencies, confirming that participants performed according to instructions and engaged in the reciprocal task. Participants sent more signals to virtual partners with lower mean response latency (i.e., more synchronous) than virtual partners with higher mean response latency. Similarly, they also responded more slowly to the more synchronous partner, which likewise suggests that participants tried to abide by the task instructions. In turn, this may indicate that participants accounted for the virtual partner’s latency to temporally align with the task. Accordingly, this reflects processes related to repetitive joint action execution, wherein individuals predict a partner’s action intention and coordinate their behavior to jointly achieve a goal (Sebanz & Knoblich, 2009).

Some studies have shown attenuated joint action execution in autism (Cerullo et al., 2021). However, many paradigms include kinematic cues that provide information about the partner’s action intentions (e.g., the shape of their hand, direction of movement). Autistic individuals may struggle to use such kinematic cues for action prediction and coordination (Gowen, 2012). Indeed, Fulceri et al. (2018) found reduced joint action coordination in autistic individuals when they only relied on kinematic cues. Conversely, Koehne et al. (2016) found comparable performance between autistic and non-autistic individuals when interacting with a virtual partner in the absence of kinematic cues. Yoo and Kim (2018) also found increased joint action coordination of drumming behaviors when accompanied by predictable timing cues (i.e., rhythmic cueing). In line with these findings, the present study only included temporal cues, wherein our participants likely relied on the rhythmic nature of the virtual partner’s response to inform their own action planning to reciprocate a response. When kinematic cues are absent and require one to exclusively rely on temporal cues, autistic and non-autistic individuals seem to use this rhythmic information in a similar way; thus, difficulties with reciprocal interactions may rather lie in the integration of social cues that require mentalization.

We further explored the neural activation patterns, regardless of synchrony, to gain insight into the underlying processes of basic reciprocal interactions. In a whole brain analysis using a pooled sample, we found expected activation in the visual and motor cortices from simply engaging in the task. We also found significant neural activation in the right IFG and extension of the motor cortex into the anterior region of the left IPL. These areas have been associated with rhythmic interactions.

Neuroimaging studies have implicated the IFG in rhythmic tapping (Kung et al., 2013) and sensorimotor synchronization with a partner (Fairhurst et al., 2013; Rao, 1997; Yun et al., 2012), demonstrating its engagement for cognitive control and adaptation. These functions are in line with the nature of the present paradigm, where participants communicated with the virtual partner at a relatively rhythmic response rate. Moreover, the IFG has also been linked to joint action execution, action observation, internally representing actions for action inference (Bolt & Loehr, 2021; Georgescu et al., 2014; Hartwright et al., 2016; Liakakis et al., 2011; Newman-Norlund et al., 2007; Ocampo et al., 2011; Su et al., 2022), as well as interpersonal awareness (Decety & Sommerville, 2003). Similarly, the IPL has been implicated in action observation, cognitive control, imitation, and self-other discrimination (Cacioppo et al., 2014; Decety & Sommerville, 2003; Gatti et al., 2017; Georgescu et al., 2014; Jackson & Decety, 2004). Accordingly, recruitment of the IFG and IPL may be necessary to engage in rhythmic reciprocal interactions that require little mentalization.

Interestingly, these neural activation patterns were comparable for autistic and non-autistic individuals, supporting the idea that reciprocal interactions that draw less on mentalization are seemingly not affected in autism. Inferring others’ actions may arguably be more straightforward for autistic individuals when the interaction partner’s behavior is more predictable. In the present paradigm, each virtual partner’s response rhythm was consistent throughout the interaction. Once participants were familiar with the tapping behavior of the respective virtual partners, they could adjust their responses without needing to infer the virtual partner’s cognitive state (e.g., intention to lead the interaction). The predictability of the interactional behavior in the present study is one possible explanation for the diminished differences observed between autistic and non-autistic participants. With recent studies showing that temporal predictability might be particularly relevant for performance in autism (Shi et al., 2022), the predictability of timing during social exchange may be crucial for understanding issues with reciprocal interactions in autism and should be investigated further.

There are limitations to consider when interpreting the results of this study. It is possible that the interactional experience of synchrony and rapport may not have been felt as strongly as was anticipated. This could be due to the minimalistic nature of the paradigm, in which little social information was provided, as well as the constraints of the fMRI environment. Yet, and in line with the original study, we used a cover story including the presence of confederates to heighten the believability of the subsequent interactions. To evaluate the effectiveness of this approach, we employed a debriefing questionnaire, in which most participants did not immediately doubt interacting with a human partner (approx. 82%). In a similar task, Bowsher-Murray et al. (2023) also suspected challenges with the interactional environment, in which they suggest that live engagement or the use of kinematic cues may boost the affiliative feeling. Moreover, regions associated with action observation and execution, but not perspective-taking and mentalization, were recruited in the present study. Thus, the presently used paradigm engaged some, but not all, regions necessary for social reciprocity. This demonstrates that processing simple and repetitive reciprocal interactions may be intact in autism, and rather points to differentiated processes when mentalization is required.

In conclusion, our findings demonstrate intact processing of basic reciprocal interactions in autism. We show similar neural and behavioral patterns for autistic and non-autistic individuals when interacting with high and low synchronous virtual partners. Atypical social reciprocity is one of the core symptoms of autism. Our findings show that this symptom is not rooted in the neural processing of and behavioral adjustment to basic rhythmic behaviors of interaction partners.

## Data and Code Availability

A preregistration, scripts, and preprocessed data are available on OSF (https://osf.io/cw7n4). Statistical maps of the fMRI analyses as well as the preprocessed behavioral ratings are provided. The full dataset underlying this article may be shared after anonymization upon reasonable request to the corresponding authors (AB, CFW). Supplementary Information are uploaded with the manuscript.

## Author Contributions

**AB:** Methodology, Software, Project administration, Investigation, Data Curation, Formal Analysis, Visualization, Writing – original draft, Writing – review & editing. **IP:** Methodology, Investigation, Supervision, Validation, Writing – review & editing. **NS:** Investigation, Writing – review & editing. **DK:** Methodology, Resources, Supervision, Writing – review & editing. **CFW:** Conceptualization, Funding Acquisition, Resources, Supervision, Writing – review & editing. **DK** and **CFW** share last authorship.

## Acknowledgements

Firstly, we would like to thank Boris Papasov for his support in setting up the MRI scanning sequences and user training. We would also like to thank the individuals who acted as confederates as part of our cover story (Marko Bierlich, Charlotte Gerard, Marie Knoke, Franz Meier, Sophia Moritz, Boris Papasov, Liisbeth Pirn, Lukas Scheel-Platz, Hannah Schupp, Hanna Späth, Marta Robles, Leora Traiger). Finally, we would like to thank our participants for their time and effort to participate in our study.

