## Supplementary Information for "Neural processing of social reciprocity in autism"

#### S1. Translated Perceived Synchrony and Rapport Questions

**Table S1.** The following questions were translated from English, as described by Cacioppo et al. (2014), into German for use in the present study that was collected from a German-speaking sample.

| English original | German translation (presently used) |
|---|---|
| 'How much rapport did you feel with your partner?' | 'Wie sympathisch war Ihnen Ihr Partner?' |
| 'How much did you trust your partner?' | 'Wie sehr haben Sie Ihrem Partner vertraut?' |
| 'How much did you like your partner?' | 'Wie sehr mochten Sie Ihren Partner?' |
| 'How synchronized was the communication between you and your partner?' | 'Wie synchron war die Kommunikation zwischen Ihnen und Ihrem Partner?' |
| 'How much would you like to work with your partner?' | 'Wie gerne würden Sie mit Ihrem Partner zusammenarbeiten?' |
| 'How much would you like to confide in your partner?' | 'Wie gerne würden Sie Ihrem Partner etwas anvertrauen?' |
| 'How close do you feel to your partner?' | 'Wie nah fühlten Sie sich Ihrem Partner?' |

#### S2. Translated Debrief Questions

**Table S2.** The following questions were developed in German and used to assess task believability in the debrief questionnaire.

| German original | English translation | Rating Labels | |
|---|---|---|---|
| 'Wie hat sich die Interaktion angefühlt?' | 'How [nice] did the interaction feel?' | 1 = sehr schlecht<br>10 = sehr gut | 1 = very bad<br>10 = very good |
| 'Wie stark hat es sich wie eine Interaktion angefühlt?' | 'How strong did it feel like an interaction?' | 1 = sehr wenig<br>10 = sehr viel | 1 = very little<br>10 = very much |

#### S3. Overview of Missing Behavioral Ratings

**Table S3.** An overview of the missing behavioral ratings for each question, including the group and condition. LL: Low latency-Low variance LH: Low latency-High variance HL: High latency-Low variance HH: High latency-High variance

| Question | Autistic | Non-autistic | Total |
|---|---|---|---|
| 'How much rapport did you feel with your partner?' | 1 | 3 | 4 |
| 'How much did you trust your partner?' | 1 | 0 | 1 |

| | | | |
|---|---|---|---|
| 'How much did you like your partner?' | 1 | 0 | 1 |
| 'How synchronized was the communication between you and your partner?' | 3 | 2 | 5 |
| 'How much would you like to work with your partner?' | 0 | 1 | 1 |
| 'How much would you like to confide in your partner?' | 1 | 1 | 2 |
| 'How close do you feel to your partner?' | 1 | 0 | 1 |

### S4. *fMRIPrep* Pre-processing Pipeline

Results included in this manuscript come from preprocessing performed using fMRIPrep 22.1.1 (Esteban, Markiewicz, et al. (2018); Esteban, Blair, et al. (2018); RRID:SCR_016216), which is based on Nipype 1.8.5 (K. Gorgolewski et al. (2011); K. J. Gorgolewski et al. (2018); RRID:SCR_002502).

#### *Preprocessing of B0 inhomogeneity mappings*

A B0-nonuniformity map (or fieldmap) was estimated based on two (or more) echo-planar imaging (EPI) references with topup (Andersson, Skare, and Ashburner (2003); FSL 6.0.5.1:57b01774).

#### *Anatomical data preprocessing*

The T1-weighted (T1w) image was corrected for intensity non-uniformity (INU) with N4BiasFieldCorrection (Tustison et al. 2010), distributed with ANTs 2.3.3 (Avants et al. 2008, RRID:SCR_004757), and used as T1w-reference throughout the workflow. The T1w-reference was then skull-stripped with a Nipype implementation of the antsBrainExtraction.sh workflow (from ANTs), using OASIS30ANTs as target template. Brain tissue segmentation of cerebrospinal fluid (CSF), white-matter (WM) and gray-matter (GM) was performed on the brain-extracted T1w using fast (FSL 6.0.5.1:57b01774, RRID:SCR_002823, Zhang, Brady, and Smith 2001). Brain surfaces were reconstructed using recon-all (FreeSurfer 7.2.0, RRID:SCR_001847, Dale, Fischl, and Sereno 1999), and the brain mask estimated previously was refined with a custom variation of the method to reconcile ANTs-derived and FreeSurfer-derived segmentations of the cortical gray-matter of Mindboggle (RRID:SCR_002438, Klein et al. 2017). Volume-based spatial normalization to two standard spaces (MNI152NLin6Asym, MNI152NLin2009cAsym) was performed through nonlinear registration with antsRegistration (ANTs 2.3.3), using brain-extracted versions of both T1w reference and the T1w template. The following templates were selected for spatial normalization: FSL's MNI ICBM 152 non-linear 6th Generation Asymmetric Average Brain Stereotaxic Registration Model [Evans et al. (2012), RRID:SCR_002823; TemplateFlow ID: MNI152NLin6Asym], ICBM 152 Nonlinear Asymmetrical template version 2009c [Fonov et al. (2009), RRID:SCR_008796; TemplateFlow ID: MNI152NLin2009cAsym].

#### *Functional data preprocessing*

For each of the 2 BOLD runs found per subject (across all tasks and sessions), the following preprocessing was performed. First, a reference volume and its skull-stripped version were generated using a custom methodology of fMRIPrep. Head-motion parameters with respect to the BOLD reference (transformation matrices, and six corresponding rotation and translation parameters) are estimated before any spatiotemporal filtering using mcflirt (FSL 6.0.5.1:57b01774, Jenkinson et al. 2002). The estimated fieldmap was then aligned with rigid-registration to the target EPI (echo-planar imaging) reference run. The field coefficients were mapped on to the reference EPI using the transform. BOLD runs were slice-time corrected to 0.991s (0.5 of slice acquisition range 0s-1.98s) using 3dTshift from AFNI (Cox and Hyde 1997, RRID:SCR_005927). The BOLD reference was then co-registered to the T1w reference using bbregister (FreeSurfer) which implements boundary-based registration (Greve and Fischl 2009). Co-registration was configured with six degrees of freedom. Several confounding time-series were calculated based on the preprocessed BOLD: framewise displacement (FD), DVARS and three region-wise global signals. FD was computed using two formulations following Power (absolute sum of relative motions, Power et al. (2014)) and Jenkinson (relative root mean square displacement between affines, Jenkinson et al. (2002)). FD and DVARS are calculated for each functional run, both using their implementations in Nipype (following the definitions by Power et al. 2014). The three global signals are extracted within the CSF, the WM, and the whole-brain masks. Additionally, a set of physiological regressors were extracted to allow for component-based noise correction (CompCor, Behzadi et al. 2007). Principal components are estimated after high-pass filtering the preprocessed BOLD time-series (using a discrete cosine filter with 128s cut-off) for the two CompCor variants: temporal (tCompCor) and anatomical (aCompCor). tCompCor components are then calculated from the top 2% variable voxels within the brain mask. For aCompCor, three probabilistic masks (CSF, WM and combined CSF+WM) are generated in anatomical space. The implementation differs from that of Behzadi et al. in that instead of eroding the masks by 2 pixels on BOLD space, a mask of pixels that likely contain a volume fraction of GM is subtracted from the aCompCor masks. This mask is obtained by dilating a GM mask extracted from the FreeSurfer's aseg segmentation, and it ensures components are not extracted from voxels containing a minimal fraction of GM. Finally, these masks are resampled into BOLD space and binarized by thresholding at 0.99 (as in the original implementation). Components are also calculated separately within the WM and CSF masks. For each CompCor decomposition, the k components with the largest singular values are retained, such that the retained components' time series are sufficient to explain 50 percent of variance across the nuisance mask (CSF, WM, combined, or temporal). The remaining components are dropped from consideration. The head-motion estimates calculated in the correction step were also placed within the corresponding confounds file. The confound time series derived from head motion estimates and global signals were expanded with the inclusion of temporal derivatives and quadratic terms for each (Satterthwaite et al. 2013). Frames that exceeded a threshold of 0.5 mm FD or 1.5 standardized DVARS were annotated as motion outliers. Additional nuisance timeseries are calculated by means of principal components analysis of the signal found within a thin band (crown) of voxels around the edge of the brain, as proposed by (Patriat, Reynolds, and Birn 2017). The BOLD time-series were resampled into standard space, generating a preprocessed BOLD run in MNI152NLin6Asym space. First, a reference volume and its skull-stripped version were generated using a custom methodology of fMRIPrep. Automatic removal of motion artifacts using independent component analysis (ICA-AROMA, Pruim et al. 2015) was performed on the preprocessed BOLD on MNI space time-series after removal of non-steady state volumes and spatial smoothing with an isotropic, Gaussian kernel of 6mm FWHM (full-width half-maximum). Corresponding "non-aggresively" denoised runs were produced after such smoothing. Additionally, the "aggressive" noise-regressors were collected and placed in the corresponding confounds file. All resamplings can be performed with a single interpolation step by composing all the pertinent transformations (i.e. head-motion transform matrices, susceptibility distortion correction when available, and co-registrations to anatomical and output spaces). Gridded (volumetric) resamplings were performed using antsApplyTransforms (ANTs), configured with Lanczos interpolation to minimize the smoothing effects of other kernels (Lanczos 1964). Non-gridded (surface) resamplings were performed using mri_vol2surf (FreeSurfer).

Many internal operations of fMRIPrep use Nilearn 0.9.1 (Abraham et al. 2014, RRID:SCR_001362), mostly within the functional processing workflow. For more details of the pipeline, see the section corresponding to workflows in fMRIPrep's documentation.

#### *Copyright Waiver*

The above boilerplate text was automatically generated by fMRIPrep with the express intention that users should copy and paste this text into their manuscripts unchanged. It is released under the CC0 license.

#### *References*

Abraham, Alexandre, Fabian Pedregosa, Michael Eickenberg, Philippe Gervais, Andreas Mueller, Jean Kossaifi, Alexandre Gramfort, Bertrand Thirion, and Gael Varoquaux. 2014. "Machine Learning for Neuroimaging with Scikit-Learn." Frontiers in Neuroinformatics 8. https://doi.org/10.3389/fninf.2014.00014.

Andersson, Jesper L. R., Stefan Skare, and John Ashburner. 2003. "How to Correct Susceptibility Distortions in Spin-Echo Echo-Planar Images: Application to Diffusion Tensor Imaging." NeuroImage 20 (2): 870–88. https://doi.org/10.1016/S1053-8119(03)00336-7.

Avants, B. B., C. L. Epstein, M. Grossman, and J. C. Gee. 2008. "Symmetric Diffeomorphic Image Registration with Cross-Correlation: Evaluating Automated Labeling of Elderly and Neurodegenerative Brain." Medical Image Analysis 12 (1): 26–41. https://doi.org/10.1016/j.media.2007.06.004.

Behzadi, Yashar, Khaled Restom, Joy Liau, and Thomas T. Liu. 2007. "A Component Based Noise Correction Method (CompCor) for BOLD and Perfusion Based fMRI." NeuroImage 37 (1): 90–101. https://doi.org/10.1016/j.neuroimage.2007.04.042.

Cox, Robert W., and James S. Hyde. 1997. "Software Tools for Analysis and Visualization of fMRI Data." NMR in Biomedicine 10 (4-5): 171–78. https://doi.org/10.1002/(SICI)1099-1492(199706/08)10:4/5<171::AID-NBM453>3.0.CO;2-L.

Dale, Anders M., Bruce Fischl, and Martin I. Sereno. 1999. "Cortical Surface-Based Analysis: I. Segmentation and Surface Reconstruction." NeuroImage 9 (2): 179–94. https://doi.org/10.1006/nimg.1998.0395.

Esteban, Oscar, Ross Blair, Christopher J. Markiewicz, Shoshana L. Berleant, Craig Moodie, Feilong Ma, Ayse Ilkay Isik, et al. 2018. "fMRIPrep 22.1.1." Software. https://doi.org/10.5281/zenodo.852659.

Esteban, Oscar, Christopher Markiewicz, Ross W Blair, Craig Moodie, Ayse Ilkay Isik, Asier Erramuzpe Aliaga, James Kent, et al. 2018. "fMRIPrep: A Robust Preprocessing Pipeline for Functional MRI." Nature Methods. https://doi.org/10.1038/s41592-018-0235-4.

Evans, AC, AL Janke, DL Collins, and S Baillet. 2012. "Brain Templates and Atlases." NeuroImage 62 (2): 911–22. https://doi.org/10.1016/j.neuroimage.2012.01.024.

Fonov, VS, AC Evans, RC McKinstry, CR Almli, and DL Collins. 2009. "Unbiased Nonlinear Average Age-Appropriate Brain Templates from Birth to Adulthood." NeuroImage 47, Supplement 1: S102. https://doi.org/10.1016/S1053-8119(09)70884-5.

Gorgolewski, K., C. D. Burns, C. Madison, D. Clark, Y. O. Halchenko, M. L. Waskom, and S. Ghosh. 2011. “Nipype: A Flexible, Lightweight and Extensible Neuroimaging Data Processing Framework in Python.” Frontiers in Neuroinformatics 5: 13. https://doi.org/10.3389/fninf.2011.00013.

Gorgolewski, Krzysztof J., Oscar Esteban, Christopher J. Markiewicz, Erik Ziegler, David Gage Ellis, Michael Philipp Notter, Dorota Jarecka, et al. 2018. “Nipype.” Software. https://doi.org/10.5281/zenodo.596855.

Greve, Douglas N, and Bruce Fischl. 2009. “Accurate and Robust Brain Image Alignment Using Boundary-Based Registration.” NeuroImage 48 (1): 63–72. https://doi.org/10.1016/j.neuroimage.2009.06.060.

Jenkinson, Mark, Peter Bannister, Michael Brady, and Stephen Smith. 2002. “Improved Optimization for the Robust and Accurate Linear Registration and Motion Correction of Brain Images.” NeuroImage 17 (2): 825–41. https://doi.org/10.1006/nimg.2002.1132.

Klein, Arno, Satrajit S. Ghosh, Forrest S. Bao, Joachim Giard, Yrjö Häme, Eliezer Stavsky, Noah Lee, et al. 2017. “Mindboggling Morphometry of Human Brains.” PLOS Computational Biology 13 (2): e1005350. https://doi.org/10.1371/journal.pcbi.1005350.

Lanczos, C. 1964. “Evaluation of Noisy Data.” Journal of the Society for Industrial and Applied Mathematics Series B Numerical Analysis 1 (1): 76–85. https://doi.org/10.1137/0701007.

Patriat, Rémi, Richard C. Reynolds, and Rasmus M. Birn. 2017. “An Improved Model of Motion-Related Signal Changes in fMRI.” NeuroImage 144, Part A (January): 74–82. https://doi.org/10.1016/j.neuroimage.2016.08.051.

Power, Jonathan D., Anish Mitra, Timothy O. Laumann, Abraham Z. Snyder, Bradley L. Schlaggar, and Steven E. Petersen. 2014. “Methods to Detect, Characterize, and Remove Motion Artifact in Resting State fMRI.” NeuroImage 84 (Supplement C): 320–41. https://doi.org/10.1016/j.neuroimage.2013.08.048.

Pruim, Raimon H. R., Maarten Mennes, Daan van Rooij, Alberto Llera, Jan K. Buitelaar, and Christian F. Beckmann. 2015. “ICA-AROMA: A Robust ICA-Based Strategy for Removing Motion Artifacts from fMRI Data.” NeuroImage 112 (Supplement C): 267–77. https://doi.org/10.1016/j.neuroimage.2015.02.064.

Satterthwaite, Theodore D., Mark A. Elliott, Raphael T. Gerraty, Kosha Ruparel, James Loughead, Monica E. Calkins, Simon B. Eickhoff, et al. 2013. “An improved framework for confound regression and filtering for control of motion artifact in the preprocessing of resting-state functional connectivity data.” NeuroImage 64 (1): 240–56. https://doi.org/10.1016/j.neuroimage.2012.08.052.

Tustison, N. J., B. B. Avants, P. A. Cook, Y. Zheng, A. Egan, P. A. Yushkevich, and J. C. Gee. 2010. “N4itk: Improved N3 Bias Correction.” IEEE Transactions on Medical Imaging 29 (6): 1310–20. https://doi.org/10.1109/TMI.2010.2046908.

Zhang, Y., M. Brady, and S. Smith. 2001. “Segmentation of Brain MR Images Through a Hidden Markov Random Field Model and the Expectation-Maximization Algorithm.” IEEE Transactions on Medical Imaging 20 (1): 45–57. https://doi.org/10.1109/42.906424.

### S5. fMRI ROI mask

**Fig S1.** The ROI mask used in the present study. The regions included are the medial superior frontal gyrus (left/right), anterior cingulate cortex (left/right), amygdala (left), parahippocampal gyrus (left), inferior parietal lobe (left), anterior gyrus (left), and the supramarginal gyrus (left).

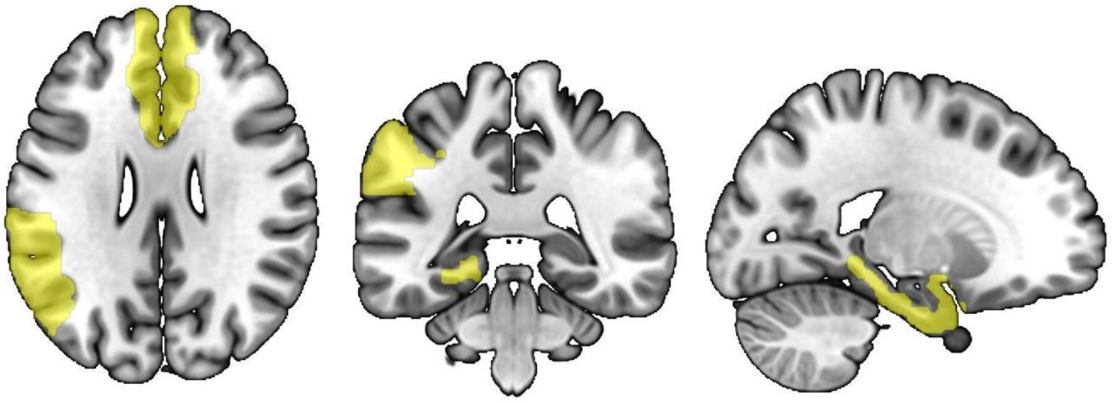


### S6. fMRI sub-analyses

#### *Replication Sub-Analysis*

In an exploratory sub-analysis, we attempted to replicate previous findings reported by Cacioppo et al. (2014) using a sub-sample including only our non-autistic participants (n = 25). The same pre-processing steps in the main text were used for the sub-analysis. At the subject-level, no parametric modulator of communication frequency was included. At the group-level, we employed a parametric, whole brain approach using FLAME1 in FSL FEAT. Cluster thresholding with a Z threshold of 2.33 and Cluster P threshold of 0.05 were used. No clusters survived for any of the contrasts of interest.

#### *Debrief Sub-Analysis*

In an exploratory sub-analysis, we checked whether the believability of the task influenced the outcome. During the debrief session, an experimenter observationally classified participants into three categories: i) did not at all suspect that they were interacting with a computer, (ii) suspected interacting with a computer only after it was specifically mentioned by the experimenter, and (iii) immediately offered their suspicions of interacting with a computer. Participants who immediately offered their suspicions of interacting with a computer were excluded for the sub-analysis (nine participants; three autistic, six non-autistic). The same pre-processing steps, subject-level contrasts, and group-level contrasts that are reported in the main text were used for the sub-analysis. The only difference is the omission of the nine participants. No clusters survived for any of the contrasts of interest.

### S7. Structural MRI comparisons

Exploratory structural analyses were conducted to compare the autistic and non-autistic groups. Using the NAMNIs pipeline (Karali et al., 2021), structural data were processed with the Juelich Brain Atlas. The intracranial volume (ICV) corrected values of grey matter and white matter for each region were compared using unpaired t-tests. No regions significantly differed, following multiple comparisons (FDR correction). The uncorrected and corrected values are reported in supplementary data file that is available on OSF: https://osf.io/cw7n4.

Karali, T., Padberg, F., Kirsch, V., Stoecklein, S., Falkai, P., & Keeser, D. (2021). NAMNIs:

Neuromodulation And Multimodal NeuroImaging software (0.3). Zenodo. https://doi.org/10.5281/zenodo.4547552

### S8. MRIqc metrics

Select metrics depict the quality of the neuroimaging data, as produced by MRIqc. Quality metrics of the functional data (Figure S1) that are reported include: DVARS rate of change of BOLD signal across each data frame (dvars_nstd), DVARS normalized with the standard deviation of the temporal difference timeseries (dvars_std), DVARS normalized with voxel-wise standard deviation of the timeseries before the temporal derivative (dvars_vstd),  mean framewise displacement (fd_mean), number of timepoints above framewise displacement threshold (fd_num), percentage of timepoints above framewise displacement threshold (fd_perc), temporal signal-to-noise ratio (tSNR). Quality metrics of the structural data (Figure S2) that are reported include: contrast-to-noise ratio (cnr), ICV fractions of the cerebral spinal fluid (icvs_csf), ICV fractions of the grey matter (icvs_gm), ICV fractions of the white matter (icvs_wm), signal-to-noise ratio (snr).

**Figure S2.** Panels depict group comparisons of select functional quality metrics.

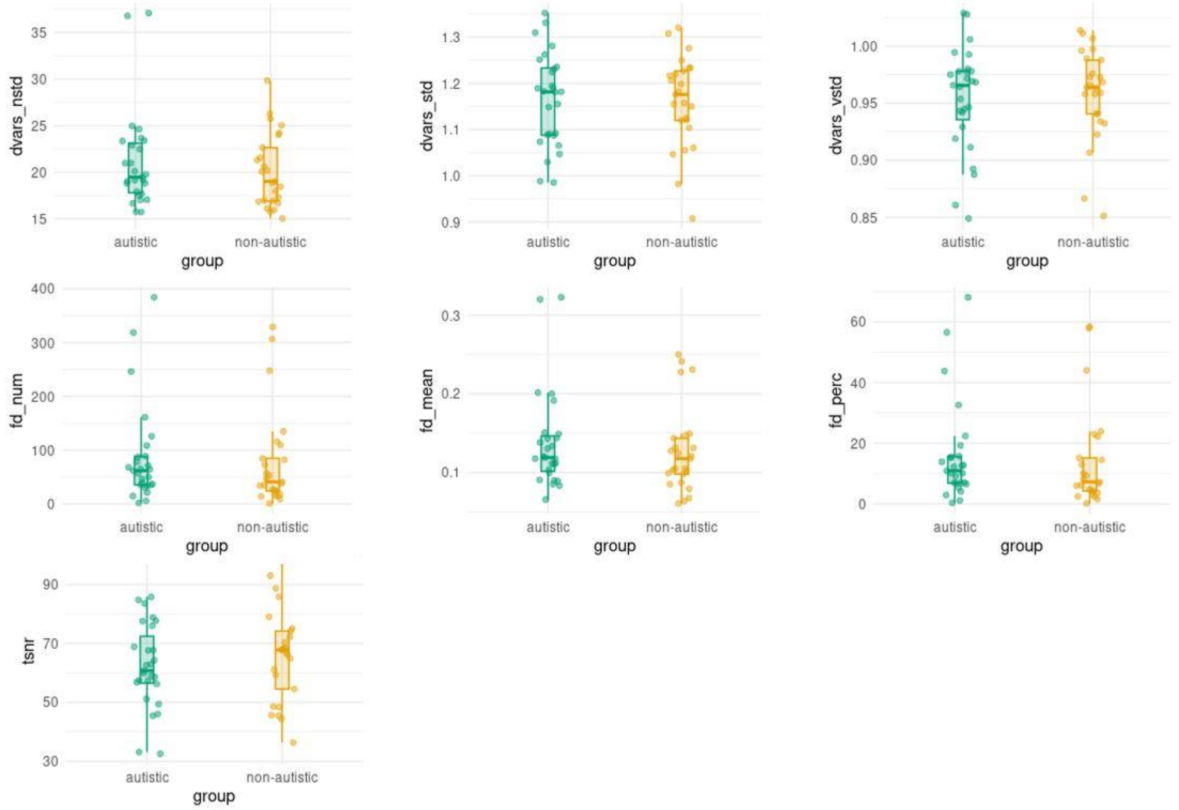

**Figure S3.** Panels depict group comparisons of select structural quality metrics.

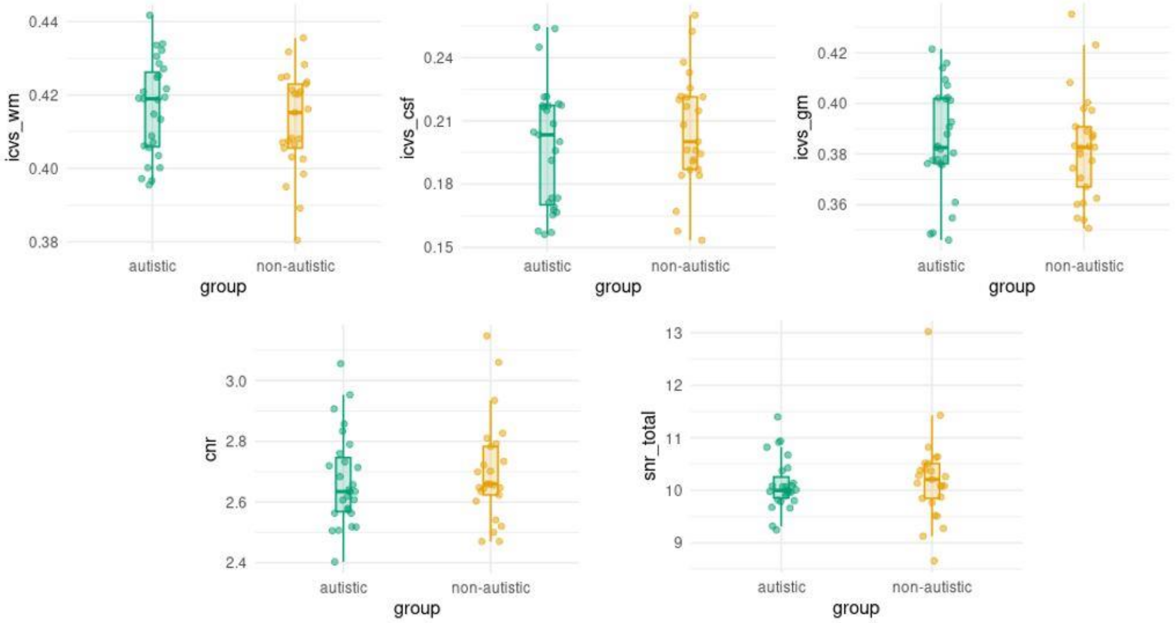

### S9. Behavioral Analyses Bayesian Mixed ANOVAs

Group: diagnostic group (non-autistic / autistic). Lat: mean latency of apparent partner's response (low / high). Var: variation of apparent partner's response (low / high).

#### *Bayesian Mixed ANOVA: perceived synchrony ratings*

**Model Comparison**

| Models | P(M) | P(M\|data) | $BF_M$ | $BF_{10}$ | error % |
|---|---|---|---|---|---|
| Null model (incl. subject and random slopes) | 0.053 | 0.378 | 10.960 | 1.000 | |
| group | 0.053 | 0.193 | 4.293 | 0.509 | 3.735 |
| lat | 0.053 | 0.131 | 2.720 | 0.347 | 1.255 |
| var | 0.053 | 0.088 | 1.734 | 0.232 | 1.586 |
| lat + group | 0.053 | 0.065 | 1.248 | 0.171 | 2.227 |
| var + group | 0.053 | 0.043 | 0.817 | 0.115 | 2.579 |
| var + lat | 0.053 | 0.031 | 0.574 | 0.082 | 2.642 |
| lat + group + lat ✻ group | 0.053 | 0.020 | 0.373 | 0.054 | 9.136 |
| var + lat + group | 0.053 | 0.014 | 0.263 | 0.038 | 1.638 |
| var + group + var ✻ group | 0.053 | 0.012 | 0.217 | 0.031 | 2.011 |

*Note.* All models include subject, and random slopes for all repeated measures factors.
*Note.* Showing the best 10 out of 19 models.

**Analysis of Effects**

| Effects | P(incl) | P(excl) | P(incl\|data) | P(excl\|data) | $BF_{incl}$ |
|---|---|---|---|---|---|
| var | 0.263 | 0.263 | 0.181 | 0.787 | 0.230 |
| lat | 0.263 | 0.263 | 0.246 | 0.714 | 0.344 |
| group | 0.263 | 0.263 | 0.320 | 0.636 | 0.502 |
| var ✻ lat | 0.263 | 0.263 | 0.014 | 0.055 | 0.257 |
| var ✻ group | 0.263 | 0.263 | 0.019 | 0.067 | 0.277 |
| lat ✻ group | 0.263 | 0.263 | 0.027 | 0.089 | 0.303 |
| var ✻ lat ✻ group | 0.053 | 0.053 | $3.783\times10^{-4}$ | $2.873\times10^{-4}$ | 1.317 |

*Note.* Compares models that contain the effect to equivalent models stripped of the effect. Higher-order interactions are excluded. Analysis suggested by Sebastiaan Mathôt.

**Fig S4. Model Averaged Q-Q Plot**

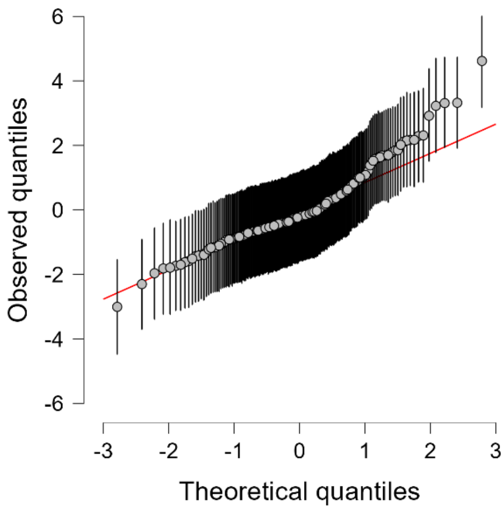

*Bayesian Mixed ANOVA: rapport ratings*

**Model Comparison**

| Models | P(M) | P(M\|data) | $BF_M$ | $BF_{10}$ | error % |
|---|---|---|---|---|---|
| Null model (incl. subject and random slopes) | 0.053 | 0.319 | 8.422 | 1.000 | |
| group | 0.053 | 0.331 | 8.923 | 1.040 | 1.365 |
| var + group | 0.053 | 0.075 | 1.451 | 0.234 | 2.678 |
| var | 0.053 | 0.068 | 1.318 | 0.214 | 1.415 |
| lat + group | 0.053 | 0.065 | 1.259 | 0.205 | 3.132 |
| lat | 0.053 | 0.059 | 1.130 | 0.185 | 0.908 |
| lat + group + lat ✻ group | 0.053 | 0.021 | 0.382 | 0.065 | 4.848 |
| var + group + var ✻ group | 0.053 | 0.018 | 0.327 | 0.056 | 2.972 |
| var + lat + group | 0.053 | 0.015 | 0.265 | 0.046 | 4.106 |
| var + lat | 0.053 | 0.012 | 0.224 | 0.039 | 1.328 |

*Note.* All models include subject, and random slopes for all repeated measures factors.

*Note.* Showing the best 10 out of 19 models.

**Analysis of Effects**

| Effects | P(incl) | P(excl) | P(incl\|data) | P(excl\|data) | BFincl |
|---|---|---|---|---|---|
| var | 0.263 | 0.263 | 0.174 | 0.795 | 0.218 |
| lat | 0.263 | 0.263 | 0.156 | 0.811 | 0.192 |
| group | 0.263 | 0.263 | 0.489 | 0.461 | 1.060 |
| var ✻ group | 0.263 | 0.263 | 0.024 | 0.097 | 0.249 |
| lat ✻ var | 0.263 | 0.263 | 0.008 | 0.036 | 0.215 |
| lat ✻ group | 0.263 | 0.263 | 0.027 | 0.088 | 0.304 |
| lat ✻ var ✻ group | 0.053 | 0.053 | 7.459×10-5 | 2.290×10-4 | 0.326 |

*Note.* Compares models that contain the effect to equivalent models stripped of the effect. Higher-order interactions are excluded. Analysis suggested by Sebastiaan Mathôt.

**Fig S5. Model Averaged Q-Q Plot**

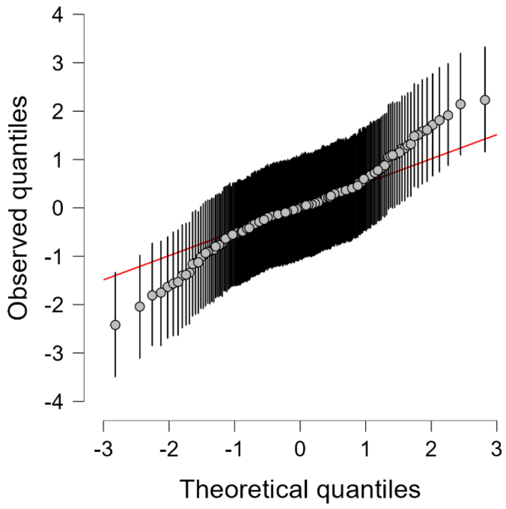

*Bayesian Mixed ANOVA: communication frequency*

**Model Comparison**

| Models | P(M) | P(M\|data) | $BF_M$ | $BF_{10}$ | error % |
|---|---|---|---|---|---|
| Null model (incl. subject and random slopes) | 0.053 | 0.010 | 0.180 | 1.000 | |
| lat | 0.053 | 0.402 | 12.123 | 40.560 | 1.567 |
| lat + group | 0.053 | 0.302 | 7.780 | 30.415 | 4.127 |
| lat + group + lat ✻ group | 0.053 | 0.078 | 1.513 | 7.815 | 4.539 |
| lat + var | 0.053 | 0.068 | 1.322 | 6.894 | 1.621 |
| lat + var + group | 0.053 | 0.056 | 1.074 | 5.673 | 7.901 |
| lat + var + group + var ✻ group | 0.053 | 0.017 | 0.309 | 1.700 | 4.045 |
| lat + var + lat ✻ var | 0.053 | 0.017 | 0.305 | 1.679 | 7.115 |
| lat + var + group + lat ✻ group | 0.053 | 0.013 | 0.233 | 1.290 | 4.822 |
| lat + var + group + lat ✻ var | 0.053 | 0.012 | 0.214 | 1.184 | 9.714 |

*Note.* All models include subject, and random slopes for all repeated measures factors.

*Note.* Showing the best 10 out of 19 models.

**Analysis of Effects**

| Effects | P(incl) | P(excl) | P(incl\|data) | P(excl\|data) | $BF_{incl}$ |
|---|---|---|---|---|---|
| lat | 0.263 | 0.263 | 0.846 | 0.021 | 39.734 |
| var | 0.263 | 0.263 | 0.141 | 0.799 | 0.176 |
| group | 0.263 | 0.263 | 0.379 | 0.499 | 0.759 |
| var ✻ lat | 0.263 | 0.263 | 0.038 | 0.159 | 0.236 |
| group ✻ lat | 0.263 | 0.263 | 0.100 | 0.391 | 0.256 |
| group ✻ var | 0.263 | 0.263 | 0.028 | 0.086 | 0.325 |
| group ✻ var ✻ lat | 0.053 | 0.053 | $3.200\times10^{-4}$ | 0.002 | 0.199 |

*Note.* Compares models that contain the effect to equivalent models stripped of the effect. Higher-order interactions are excluded. Analysis suggested by Sebastiaan Mathôt.

**Fig S6. Model Averaged Q-Q Plot**

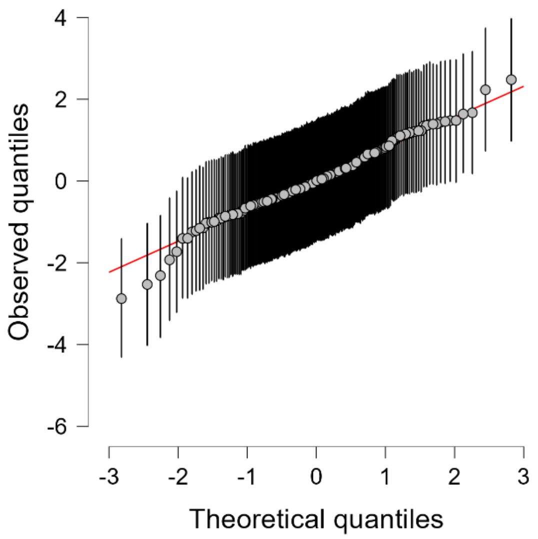

*Bayesian Mixed ANOVA: participants' average response latency*

**Model Comparison**

| Models | P(M) | P(M\|data) | $BF_M$ | $BF_{10}$ | error % |
|---|---|---|---|---|---|
| Null model (incl. subject and random slopes) | 0.053 | 0.018 | 0.329 | 1.000 | |
| lat | 0.053 | 0.268 | 6.575 | 14.886 | 5.448 |
| lat + group | 0.053 | 0.217 | 4.976 | 12.050 | 8.533 |
| var + lat + group + var ✻ group | 0.053 | 0.093 | 1.856 | 5.202 | 7.304 |
| var + lat | 0.053 | 0.092 | 1.823 | 5.116 | 9.687 |
| var + lat + group | 0.053 | 0.061 | 1.179 | 3.421 | 5.388 |
| lat + group + lat ✻ group | 0.053 | 0.056 | 1.074 | 3.133 | 7.228 |
| var + lat + group + var ✻ lat + var ✻ group | 0.053 | 0.041 | 0.761 | 2.256 | 6.461 |
| var + lat + var ✻ lat | 0.053 | 0.032 | 0.587 | 1.756 | 1.919 |
| var + lat + group + var ✻ group + lat ✻ group | 0.053 | 0.028 | 0.512 | 1.538 | 7.134 |

*Note.* All models include subject, and random slopes for all repeated measures factors.
*Note.* Showing the best 10 out of 19 models.

**Analysis of Effects**

| Effects | P(incl) | P(excl) | P(incl\|data) | P(excl\|data) | $BF_{incl}$ |
|---|---|---|---|---|---|
| var | 0.263 | 0.263 | 0.182 | 0.572 | 0.319 |
| lat | 0.263 | 0.263 | 0.731 | 0.048 | 15.288 |
| group | 0.263 | 0.263 | 0.321 | 0.415 | 0.775 |
| var ✻ lat | 0.263 | 0.263 | 0.114 | 0.293 | 0.389 |
| var ✻ group | 0.263 | 0.263 | 0.178 | 0.117 | 1.512 |
| lat ✻ group | 0.263 | 0.263 | 0.119 | 0.438 | 0.272 |
| var ✻ lat ✻ group | 0.053 | 0.053 | 0.004 | 0.009 | 0.448 |

*Note.* Compares models that contain the effect to equivalent models stripped of the effect. Higher-order interactions are excluded. Analysis suggested by Sebastiaan Mathôt.

**Fig S7. Model Averaged Q-Q Plot**

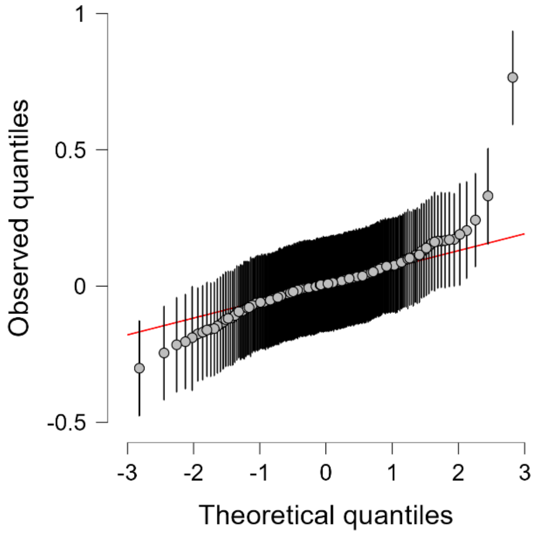

### S10. Additional visualizations for the effect of task

**Fig S8.** The surviving clusters on the t-statistic map for the effect of task after correction with TFCE thresholding overlaid on the uncorrected t-statistic map.

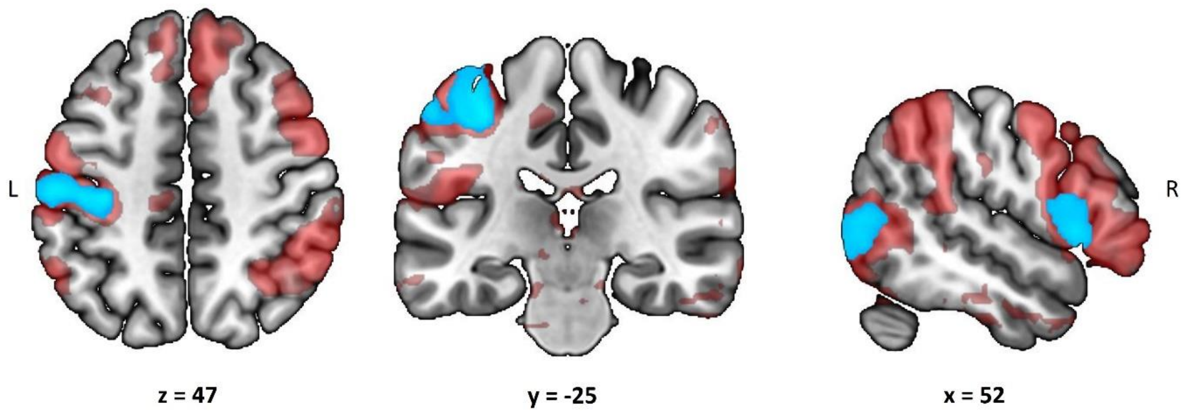
